# Predictive Modeling and Deep Phenotyping of Obstructive Sleep Apnea and Associated Comorbidities through Natural Language Processing and Large Language Models

**DOI:** 10.1101/2024.04.19.24306084

**Authors:** Awwal Ahmed, Anthony Rispoli, Carrie Wasieloski, Ifrah Khurram, Rafael Zamora-Resendiz, Destinee Morrow, Aijuan Dong, Silvia Crivelli

## Abstract

Obstructive Sleep Apnea (OSA) is a prevalent sleep disorder associated with serious health conditions. This project utilized large language models (LLMs) to develop lexicons for OSA sub-phenotypes. Our study found that LLMs can identify informative lexicons for OSA sub-phenotyping in simple patient cohorts, achieving wAUC scores of 0.9 or slightly higher. Among the six models studied, BioClinical BERT and BlueBERT outperformed the rest. Additionally, the developed lexicons exhibited some utility in predicting mortality risk (wAUC score of 0.86) and hospital readmission (wAUC score of 0.72). This work demonstrates the potential benefits of incorporating LLMs into healthcare.

**Data and Code Availability:** This paper uses the MIMIC-IV dataset (Johnson et al., 2023a), which is available on the PhysioNet repository (Johnson et al., 2023b). We plan to make the source code publicly available in the future.

**Institutional Review Board (IRB):** This research does not require IRB approval.

## 1. Introduction

Obstructive sleep apnea (OSA) is a sleep-related breathing disorder characterized by complete (apnea) or partial (hypopnea) obstruction of the upper airway, leading to sleep disruptions and intermittent hypoxemia, i.e., a low level of oxygen in the blood. In the United States, OSA prevalence among adults is estimated at 12% (Watson, 2016), and almost one billion people are affected globally (Benjafield et al., 2019). Additionally, OSA is linked to an increased risk of various cardiovascular (CV)(Loke et al., 2012; Seiler et al., 2019; Sulit et al., 2006; Xia et al., 2018), metabolic(Sulit et al., 2006), and neurological conditions (Olaithe et al., 2018; Yaffe et al., 2011).

While previous studies indicate some correlation between OSA severity and susceptibility to cardiovascular and metabolic diseases, the pervasive and often undetected nature of OSA, coupled with the challenge of identifying individuals most at risk for CV and metabolic dysfunction, has resulted in under-diagnosis and consequently suboptimal healthcare utilization and poor personalized patient care. In 2015, the U.S. spent $12.4 billion on OSA-related expenses, with substantial costs in diagnosis, treatment, and associated productivity losses. Furthermore, the report estimated the cost burden of undiagnosed OSA among U.S. adults was an astounding $149.6 billion during the same period (Watson, 2016).

Diagnosing OSA is complex and involves specialized tests like polysomnography (PSG) and home sleep apnea testing (HSAT). However, research indicates that the likelihood of misdiagnosis in OSA cases based on a single night ranges from 20% to 50% (Lechat et al., 2022). There is an urgent need to improve OSA deep phenotyping and patient risk assessment to mitigate the risk of developing multiple highly challenging health conditions. Deep phenotyping is defined as the precise and comprehensive analysis of phenotypic abnormalities for scientific examination of human disease (Robinson, 2012).

The widespread adoption of electronic health records (EHRs) has increased the availability and quality of electronic medical data. Utilizing EHRs, mainly the unstructured patient discharge notes, this study seeks to employ natural language processing (NLP) and large language models (LLMs) for a comprehensive understanding of obstructive sleep apnea and associated comorbidities. The three primary objectives are as follows:

- Compare LLMs: Compare large language models to evaluate their effectiveness in extracting informative n-grams for characterizing obstructive sleep apnea and its associated comorbidities.
- Develop Predictive Models for Diagnosis: Develop predictive models to sub-phenotype and diagnose obstructive sleep apnea in diverse patient cohorts. This involves utilizing unstructured discharge notes annotated with LLM-selected n-grams for diagnosis.
- Forecast Medical Outcomes: Construct predictive models with historical EHR data to forecast future medical outcomes, with a specific focus on predicting patient mortality rates and hospital readmissions. The overarching goal is to contribute to improved clinical decision-making.

## 2. Related Work

### 2.1. Large Language Models in Health Care

Large language models (LLMs) are a type of artificial intelligence (AI) model that is trained on vast amounts of typically unlabeled data. While traditional AI models are often single-task systems, LLMs, capable of performing many different downstream tasks after training, represent a paradigm shift in AI model development (Bommasani et al., 2021). This allows a single LLM to be reused across a range of tasks with minimal adaptation or retraining. However, LLMs typically have a substantially greater number of parameters than traditional AI models—sometimes in the hundreds of billions. This requires significant computational resources for training (Le Scao et al., 2022).

Recent advancements in LLMs, the exponential growth of medical literature, and the widespread availability of large-scale EHRs have set the stage for clinical LLMs to revolutionize medical practice.

Noteworthy applications of LLMs in healthcare include named entity recognition and relation extraction (e.g., BioBert by Lee et al. (2020), and BlueBert by Peng et al. (2019)), medical question answering and inference (e.g., GatorTron by Yang et al. (2022), and Med-PaLM by Singhal et al. (2023)), discharge summaries (e.g., ChatGPT by Patel and Lam (2023)), diagnosis classification (e.g., Clinical-Bert by Alsentzer et al. (2019)), and various others (Luo et al., 2022; Yang et al., 2022).

With LLMs, embeddings are numerical representations of words, phrases, or sentences that capture contextual information and understand relationships within large segments of text. They have been employed in various tasks, such as text retrieval and ranking (e.g., Qadrud-Din et al. (2020)), text classification (e.g.,Chae and Davidson (2023)), and sentiment analysis (e.g., Savelka and Ashley (2023)). In this project, our focus lies in embeddings extracted from LLMs. Given a set of initial medical terms (referred to as seed terms) for categorizing OSA and associated comorbidities, we are interested in expanding the lexicon through the LLMs, i.e., searching for similar medical terms by computing the cosine similarity between embeddings.

### 2.2. Predictive Modeling for the Diagnosis of OSA and Associated Comorbidities

The in-lab polysomnography (PSG) is the gold standard test to diagnose OSA, but with significant cost. The Home Sleep Apnea Test (HSAT) is an acceptable alternative when PSG is not feasible. However, HSAT is not appropriate in patients with severe pulmonary disease, congestive heart failure, neuromuscular disease, or certain other sleep disorders (Goyal and Johnson, 2017). OSA is widely underdiagnosed—86% to 95% of individuals found in population surveys with clinically significant obstructive sleep apnea syndrome report no prior diagnosis (Randerath et al., 2021).

Coupled with machine learning (ML) and natural language processing (NLP), data collected as part of routine sleep clinic visits, both structured (such as demographics, medications, procedures, and lab results) and unstructured (such as clinical notes and diagnostic images), can be repurposed to improve sleep phenotyping accuracy and investigate the relationship to comorbidities (Cade et al., 2022; Keenan et al., 2020; Ramesh et al., 2021; Strausz et al., 2020).

EHRs present a pragmatic pathway for understanding OSA on a large scale. While structured clinical data (such as demographics, medications, procedures, imaging, and lab results) has been widely used in research to improve OSA diagnosis and clinical efficiency, a significant amount of rich, fine-grained patient information is embedded in unstructured clinical notes. In this project, our focus is on developing predictive models to sub-phenotype and diagnose obstructive sleep apnea across diverse patient cohorts. To achieve this, we utilize LLM-expanded medical terms as features for our predictive models.

### 2.3. Predicting Mortality and Hospital Readmission through NLP Techniques

Recent studies have explored the application of NLP techniques to predict mortality and hospital readmission in healthcare settings. Some approaches used unstructured clinical notes only (Boag et al., 2018; Huang et al., 2019; Wu et al., 2023; Ye et al., 2020). For example, Huang et al. pre-trained BERT on clinical notes and fine-tuned it for improved 30-day hospital readmission prediction.

Yet, others integrate clinical text, vital signs, time series measurements, and imaging to create a comprehensive profile of a patient’s health status for more accurate predictions (Ashfaq et al., 2019; Chen et al., 2022; Jin et al., 2018; Khadanga et al., 2019; Parreco et al., 2018). For example, Jin et al. performed named entity extraction and negation detection on clinical notes and trained a multimodal neural network that integrated time series signals and unstructured clinical text representations for predicting in-hospital mortality risk in ICU patients.

In this study, we aim to assess the predictive effectiveness of LLM-expanded medical terms, designed for OSA diagnosis, in predicting mortality and hospital readmission risks.

## 3. Methodology

### 3.1. Dataset

The Medical Information Mart for Intensive Care (MIMIC)-IV database is utilized for this project (Johnson et al., 2023b). It comprises deidentified electronic health records for patients admitted to the Beth Israel Deaconess Medical Center emergency department or ICU between 2008 and 2019. MIMIC-IV v2.2, released in January 2023, consists of 299,712 patients and 431,231 admissions.

In addition to obstructive sleep apnea, we examined the following associated comorbidities: diabetes mellitus type 2 (T2DM), hypertension (HTN), heart failure (HF), and atrial fibrillation (AF). Physicians have selected a specific set of International Classification of Diseases (ICD) codes for each health condition, and you can find a summary in Table 1.

**Table 1:** Summary of ICD Codes and Seed Terms.

| | ICDs, $N$ | Seed Terms, $N$ |
| --- | --- | --- |
| OSA | 9 | 38 |
| T2DM | 157 | 13 |
| HTN | 68 | 10 |
| HF | 61 | 15 |
| AF | 16 | 12 |
| Total | 311 | 88 |

As an example, the following ICD codes are used to identify patients with OSA: 327.20 (Organic sleep apnea, unspecified), 327.23 (Obstructive sleep apnea [adult, pediatric]), 327.29 (Other organic sleep apnea), 780.51 (Insomnia with sleep apnea), 780.53 (Hypersomnia with sleep apnea), 780.57 (Sleep apnea [NOS]), G4730 (Sleep apnea, unspecified), G4733 (Obstructive sleep apnea [adult, pediatric]), and G4739 (Other sleep apnea). A patient is considered to have a positive diagnosis for a specific health condition if they possess at least one corresponding ICD code. Table 2 provides basic demographic data of patients of interest.

**Table 2:** Patient Demographics.

|  | OSA | T2DM | HTN | HF | AF |
| --- | --- | --- | --- | --- | --- |
| Patients, $N$ | 13,942 | 21,666 | 74,080 | 21,076 | 25,743 |
| Discharge Notes, $N$ | 29,892 | 53,446 | 161,245 | 49,479 | 55,418 |
| Women, $N(\%)$ | 5,628 (40.4) | 10,088 (46.6) | 36,486 (49.3) | 9,944 (47.2) | 11,215 (44.0) |
| White, $N(\%)$ | 9,895 (71.0) | 13,802 (63.7) | 51,238 (69.2) | 15,269 (72.4) | 19,980 (77.6) |
| Black, $N(\%)$ | 1,966 (14.1) | 3,688 (17.0) | 9,959 (13.4) | 2,476 (11.7) | 1,776 (6.9) |
| Other, $N(\%)$ | 2,081 (14.9) | 4,176 (19.3) | 12,883 (17.4) | 3,331 (15.9) | 3,967 (15.5) |

**Table 3:**
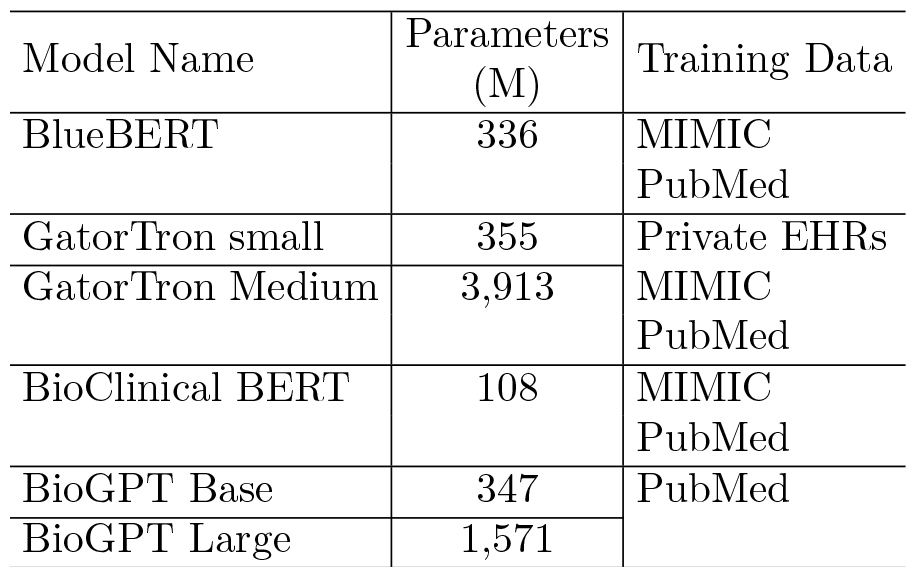
Summary of LLMs.

### 3.2. Process Flow

The flowchart (Figure 1) represents a data-driven approach to healthcare analysis and it outlines the sequence of tasks or processes that are executed to accomplish the three objectives, as described in Section 1, Introduction.

**Figure 1.**
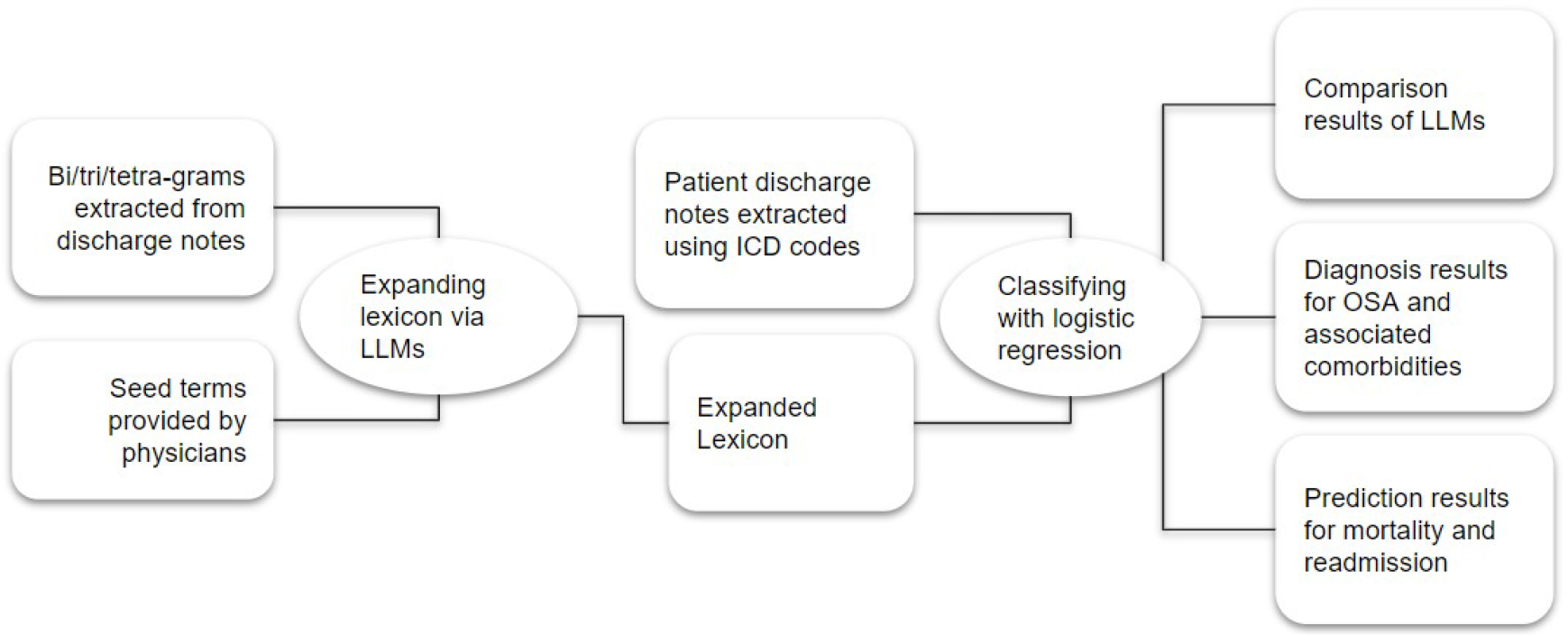
A Step-by-step Process Flow

#### Bi/Tri/Tetra-grams extracted from discharge notes

The initial step is to extract bigrams (pairs of consecutive words), trigrams (triplets of consecutive words), and tetragrams (four consecutive words) from patient discharge notes to capture commonly used phrases. MIMIC-IV v2.2 contains 331,794 discharge notes. The mean number of characters per note is 10,551. The longest and shortest discharge notes have 60,381 and 353 characters, respectively. The numbers of bigrams, trigrams, and tetragrams extracted from this step are 3,096,096, 5,407,839, and 4,792,806, respectively. These ngrams (i.e., bigrams, trigrams, and tetragrams) are candidates for expanding lexicons in this study.

#### Seed terms provided by physicians

Physicians involved in this study provided seed terms, relevant medical terms or phrases for OSA and comorbidities of interest. The number of seed terms per condition is listed in Table 1. As an example, the following terms are among the 38 terms for OSA: poorly refreshing sleep, obstructive sleep apnea (OSA), obesity hypoventilation syndrome (OHS), unrefreshing sleep, sleepiness, excessive daytime sleepiness (EDS), and snoring.

#### Expanding lexicon via LLMs

The extracted ngrams are used to expand the seed terms using LLMs. The goal of this step is to identify informative ngrams (bi/tri/tetra-grams) from discharge notes for categorizing OSA and its associated comorbidities. The general approach to selection is by comparing how similar these ngrams are to the seed terms of corresponding conditions. The similarity between an ngram and a seed term is measured by the cosine similarity between their embeddings, which are semantic representations extracted from an LLM. Specifically, given an ngram *t* and a seed term *s*, the cosine similarity between *t* and *s* is measured using

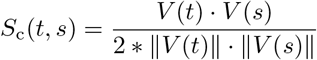

where *V* (*t*) is the LLM embedding vector for the ngram and *V* (*s*) is the LLM embedding vector for the seed term. We investigated multiple LLMs and compared their representation power or quality.

#### Expanded Lexicon

For each ngram, there are 88 similarity scores, corresponding to the 88 seed terms (Table 1). The importance or relevance of each ngram to a specific health condition (OSA or comorbidity) is measured by the average of all similarity scores between the ngram and the seed terms associated with that condition. As a result, each ngram has five similarity scores, one for each health condition.

The similarity scores of ngrams are then ranked individually for each condition, and the rankings of bigrams, trigrams, and tetragrams are separated as well. Thus, each health condition ends with three distinct ranked lists, from which a number of ngrams are selected as textual features to be used for classification.

#### Patient discharge notes extracted using ICD codes

Discharge notes for patients with OSA and/or comorbidities are extracted based on ICD codes (See Table 2 for summary). The process involved merging information from multiple tables or files. Discharge notes are long-form narratives that describe the reason for a patient’s admission to the hospital, their hospital course, and any relevant discharge instructions. Each discharge note is for a patient’s one hospital stay, and a patient may have multiple discharge notes if he or she has more than one hospital stay.

#### Classifying with logistic regression

Multinomial Logistic Regression (MLR) is a statistical technique in machine learning designed to model relationships among multiple categories or classes within a dependent variable. MLR estimates probabilities for each class and is selected for this study due to its simplicity and its ability to offer valuable insights into the relative importance of different text features (i.e., ngrams) for predicting outcomes.

Each discharge note was labeled based on the diagnosis or medical outcome. For instance, in a study involving patients with OSA and HF (Table 4), the labels include OSA only (i.e., without HF), HF only (i.e., without OSA), and OSA & HF (i.e., with both OSA and HF). For the mortality study, each discharge note was labeled with either alive or deceased.

**Table 4:** Patient Cohort for OSA and HF.

| Diagnosis Label | Discharge Notes (N) |
| --- | --- |
| OSA Only (w/o HF) | 21,929 |
| HF Only (w/o OSA) | 41,516 |
| OSA & HF | 7,963 |
| Total | 71,408 |

To represent each note, a “bag-of-ngrams” encoding was applied, treating each ngram as a presence/absence feature variable. The selection of ngrams is determined by the health conditions under study. For instance, in a study involving patients with OSA and HF using trigrams, the trigrams may comprise the top-n trigrams from OSA’s trigram list and the top-n trigrams from HF’s trigram list. These two listed are then merged with duplicates removed.

Models were tested along a repeated stratified 10-fold validation scheme.

#### Comparison results of LLMs

LLMs represent ngrams differently. To assess the effectiveness of the LLMs in expanding the lexicon and aiding in accurately characterizing obstructive sleep apnea and its associated comorbidities, diagnosis performance is used. See Section 4 for details.

#### Diagnosis results for OSA and associated comorbidities

Diverse patient cohorts were investigated with different approaches. See Section 5 for details.

#### Prediction results for mortality and readmission

Although seed terms and then expanded lexicon are for characterizing OSA and associated comorbidities, the predictive power in mortality and readmission risks were assessed in Section 6.

### 3.3. Software and Platform

The packages used in this study include pandas, NumPy, multiple large language models, Matplotlib, NLTK, mpi4py, PyTorch, and scikit-learn. The National Energy Research Scientific Computing Center (NERSC) available at the Lawrence Berkeley National Laboratory provided computing resources used for LLMs, complex machine learning models and processing a large volume of electronic health records.

## 4. Comparative Study of Large Language Models

The representation power of large language models depends on their model size, exposure to training data, and the ability to capture intricate linguistic patterns and contextual dependencies. In this section, our objective is to compare the ability of six large language models to identify and discover informative ngrams from clinical notes for categorizing OSA and associated comorbidities. Table 3 shows how these models differ in size, training data, and model architecture.

### 4.1. Comparing LLMs in the Diagnosis of Sleep Apnea and Heart Failure

The goal is to compare the abilities of the six large language models to identify trigrams for diagnosing patients with obstructive sleep apnea, heart failure, or both. Table 4 provides information on the patient cohort composition and the number of discharge notes in each group.

Building on our previous study with MIMIC-III, trigrams were used in this study. An equal number of trigrams were chosen from the ranked trigram lists of OSA and HF. If a trigram is relevant to both conditions and appears in both lists, only one copy is kept when merging the two lists. After merging, the actual counts of unique trigrams vary across LLMs (Table 5). Six trigram or feature sizes were explored.

**Table 5:** Summary of Trigram Counts across LLMs.

| Trigram Count<br>(Before Dropping<br>Duplicates) | Trigram Count (After Dropping Duplicates) |  |  |  |  |  |
| --- | --- | --- | --- | --- | --- | --- |
|  | BlueBERT | GatorTron<br>Small | GatorTron<br>Medium | BioClinical BERT | BioGPT<br>Base | BioGPT<br>Large |
| 50,000 | 47,793 | 36,373 | 28,186 | 46,673 | 31,974 | 30,049 |
| 100,000 | 80,861 | 69,246 | 55,920 | 90,199 | 59,995 | 57,171 |
| 150,000 | 116,637 | 100,927 | 82,982 | 131,812 | 87,052 | 83,608 |
| 200,000 | 150,705 | 132,099 | 109,730 | 172,136 | 113,390 | 109,788 |
| 250,000 | 183,595 | 162,900 | 135,896 | 211,506 | 139,231 | 135,692 |
| 300,000 | 215,398 | 193,201 | 161,894 | 249,923 | 165,236 | 161,415 |
| 350,000 | 246,587 | 223,121 | 187,600 | 287,594 | 191,102 | 187,492 |

Each selected trigram serves as a unique feature for the bag-of-words classification model, capturing the presence or absence of each trigram in a given discharge note. The discharge notes are labeled with OSA only (i.e., without HF), HF only (i.e., without OSA), or both OSA & HF based on the ICD code. Multinomial logistic regression (MLR) was then employed for a 3-class 1-to-many classification. The classification performance, represented by wAUC (weighted area under the curve), indicates the quality of trigrams selected by LLMs. In other words, it offers insights into how effectively the chosen trigrams contribute to the model’s ability to accurately classify patients into their respective diagnostic categories.

Each line in Figure 2 displays a trend of increasing wAUC score with the rise in trigram count, suggesting that model performance generally improves with the utilization of more trigrams or features. The actual number of trigrams (i.e., after duplicates are removed) ranges from 28,186, which is 0.52% of all trigrams, to 287,594, constituting 5.3% of all trigrams. The wAUC score spans from approximately 0.72 to 0.86.

**Figure 2.**
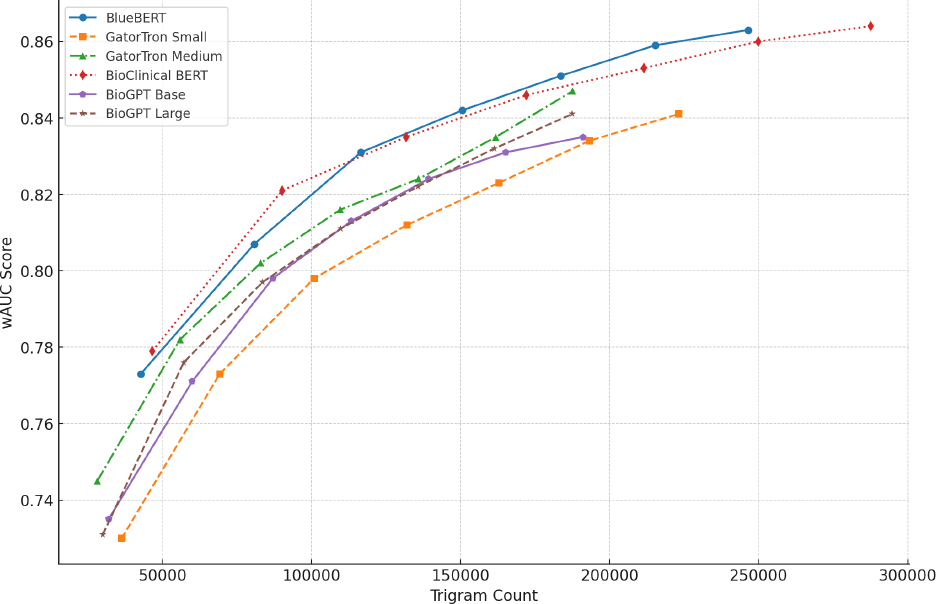
LLM Performance with Varying Trigram Counts

According to the trends, BlueBERT and BioClinical BERT demonstrate better performance, especially at higher trigram counts. Table 5 reveals that these models have larger trigram counts post-deduplication than other large language models, suggesting that their OSA and HF trigram lists are more selective, that is, they share fewer trigrams. Furthermore, BioGPT Large outperforms BioGPT Base and GatorTron Medium outperforms GatorTron Small, potentially indicating that the more complex models are selecting trigrams of higher quality, thereby better characterizing the two health conditions.

### 4.2. Comparing the Impact of Ngram Size

The goal of this study is to evaluate the effectiveness of different ngram sizes (i.e., bigram, trigram, and tetragram) in terms of diagnostic accuracy, as measured by the wAUC scores. The same patient cohort (Table 4) as that of Section 4.1 and BioClinical BERT were used in this study.

Similar to that of Section 4.1, an equal number of bigrams, trigrams, or tetragrams were chosen from the OSA and HF lists. Since a ngram can be relevant to both conditions, the actual count of unique ngrams, after combining the two lists, varies. Six ngram or feature sizes were explored, Overall, Figure 3 shows the wAUC range from approximately 0.74 to 0.9. It also suggests that the wAUC Score for all three types of ngrams increases as the number of ngrams grows, with the bigram model generally outperforming trigram and tetragram models, indicating that the additional context provided by trigrams and tetragrams does not contribute to a higher predictive performance for this analysis.

**Figure 3.**
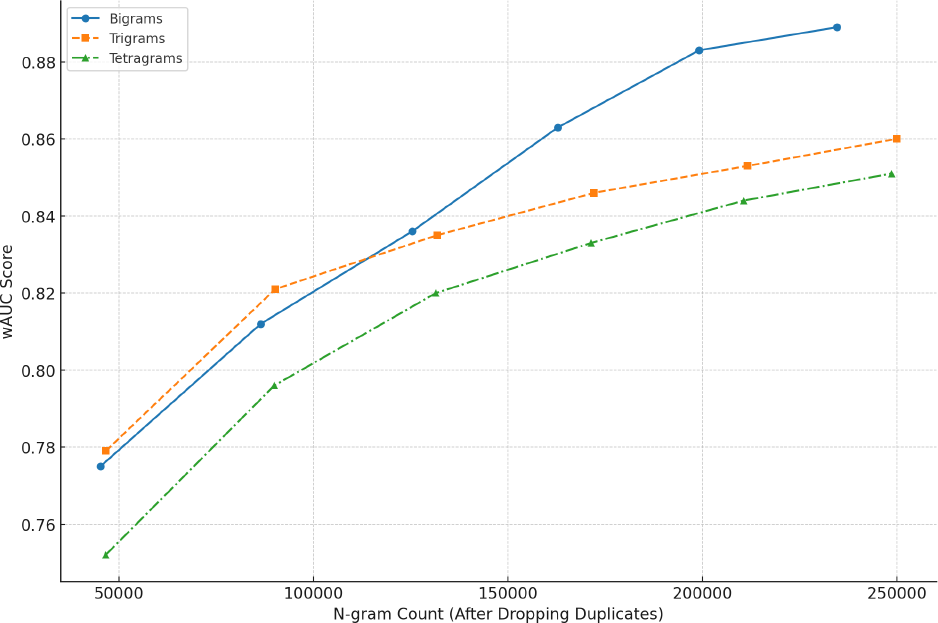
LLM Performance with Different N-gram Models

## 5. Predictive Modeling for Sleep Apnea Diagnosis

Sleep apnea is associated with increased cardiovascular and metabolic risks. We aim to evaluate how informative LLM-selected lexicons are in distinguishing OSA from common comorbidities. Section 4 used patients with OSA and/or HF for LLMs comparison; this section addressed other common comorbidities, including OSA and type 2 diabetes mellitus (T2DM), OSA and hypertension (HTN), and OSA and Atrial Fibrillation (AF) (Table 6).

**Table 6:** Summary of Three Patient Cohorts.

| OSA and T2DM Cohort |  | OSA and HTN Cohort |  | OSA and AF Cohort |  |
| --- | --- | --- | --- | --- | --- |
| Diagnostic Label | Discharge Notes(N) | Diagnostic Label | Discharge Notes (N) | Diagnostic Label | Discharge Notes(N) |
| OSA only (w/o T2DM) | 22,624 | OSA Only (w/o HTN) | 12,534 | OSA Only (w/o AF) | 22,698 |
| T2DM Only (w/o OSA) | 46,178 | HTN Only (w/o OSA) | 55,000 | AF Only (w/o OSA) | 48,224 |
| OSA & T2DM | 7,268 | OSA & HTN | 17,358 | OSA & AF | 7,194 |
| **Total** | **76,070** |  | **84,892** |  | **78,116** |

Trigrams were used in the OSA and T2DM, and OSA and HTN cohorts, while tetragrams were used in the OSA and AF cohort. Two LLMs—BlueBERT and GatorTron Medium—were used. For each patient cohort, an equal number of LLM-selected trigrams or tetragrams from both obstructive sleep apnea (OSA) and the corresponding comorbidity in the cohort were merged. Each unique trigram or tetragram functioned as a presence/absence feature variable for encoding discharge notes. Multinomial logistic regression was again employed for a 3-class 1-to-many classification. A search for hyperparameters, such as regularization strength and different optimization solvers, was conducted. All models underwent testing using a repeated stratified 10-fold cross-validation approach. The performance measures in Table 7 are wAUC scores.

**Table 7:**
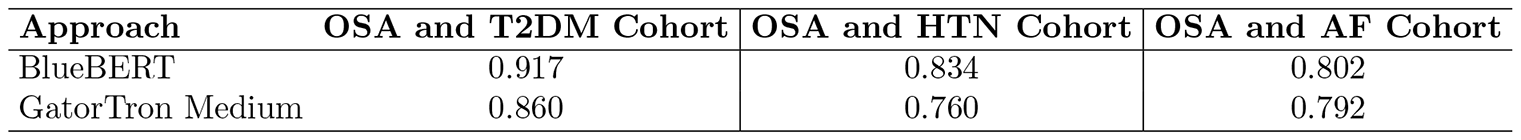
Diagnostic Results for the Three Patient Cohorts.

| Approach | OSA and T2DM Cohort | OSA and HTN Cohort | OSA and AF Cohort |
| --- | --- | --- | --- |
| BlueBERT | 0.917 | 0.834 | 0.802 |
| GatorTron Medium | 0.860 | 0.760 | 0.792 |

Table 7 shows BlueBERT consistently outperforms GatorTron Medium in all cohorts, with wAUC ranging from 0.917 for the OSA and T2DM cohort and 0.802 for the OSA and AF cohort. Additionally, based on Section 4.2, trigram model performed slightly better than tetragam model, this might be contributed to the performance variations between trigrams (OSA and T2DM Cohort and OSA and HTN Cohort) and tetragrams (OSA and AF Cohort).

To further investigate the impact of LLM-selected ngrams on OSA diagnosis, we included a generic ngram model, i.e., the Top-ngram approach in Table 8. Instead of using LLM-selected trigrams, this approach uses the most frequently appearing trigrams in discharge notes from the MIMIC-IV dataset. Table 8 shows that the Top-ngram approach yields comparable or better results for OSA diagnosis across three patient cohorts.

**Table 8:**
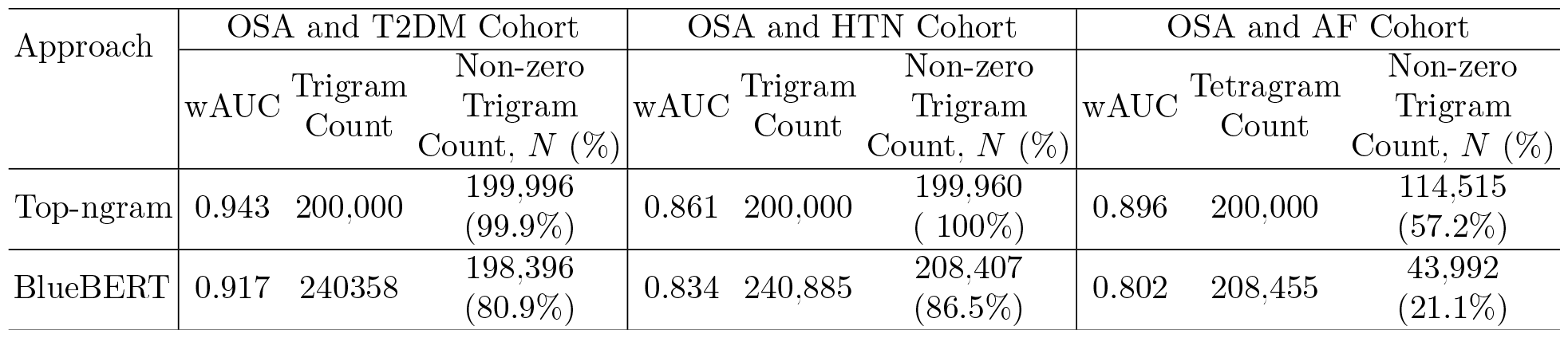
Comparing Diagnostic Results: Top-ngram vs. BlueBERT.

We then examined the model parameters for a better understanding of the models. Specifically, we analyzed the number of unique ngrams used in each model (i.e., Trigram Count) and the number of grams with non-zero coefficients in the fitted MLR models (i.e., Non-zero Trigram Count)(Table 8). Please note that a non-zero coefficient indicates that an ngram contributes to the model performance. Table 8 shows that the Top-ngram approach utilized nearly all of the initial 200,000 ngrams, while BlueBERT started with a larger pool of trigrams; however, a significantly lower percentage had non-zero coefficients, suggesting a more sparse model. This is often desirable as it indicates a more parsimonious representation, potentially enhancing model interpretability and reducing the risk of overfitting to noise in the data. Preliminary studies also indicate that the BlueBERT model runs about 5 times faster than the Top-ngram approach.

## 6. Predicting Mortality and Hospital Readmission

This study aims to construct predictive models using historical EHR data to forecast future medical outcomes. Our focus is on evaluating the predictive efficacy of LLM-expanded lexicon, designed for OSA diagnosis, in predicting mortality and hospital readmissions.

### 6.1. Mortality Prediction

We extracted patients with obstructive sleep apnea (OSA), type 2 diabetes mellitus (T2DM), or hypertension (HTN). Patients’ discharge notes were labelled as deceased if patients expired either within 6 months after discharge or one year after discharge. The numbers in Table 9 are discharge note counts for different groups Two approaches were explored. The Top-ngram approach used the top 200,000 most frequently appearing trigrams in discharge notes, while the GatorTron Medium approach merged the top trigrams from each health condition’s GatorTron-expanded trigram lists, resulting in a final trigram count of 264,857.

**Table 9:** The Mortality Study Cohort.

|  | 6-month Post-discharge | 1-year Post-discharge |
| --- | --- | --- |
| Alive | 14,000 | 20,000 |
| Deceased | 9,606 | 14,796 |

Table 10 shows that the Top-ngram approach out-performed the GatorTron model in both 6-month and 1-year mortality predications, suggesting that lexicons identified by LLMs have some but limited utility in predicting mortality risk. Yet, the GatorTron model had a much lower non-zero trigram feature, suggesting a simpler model.

**Table 10:** Mortality Prediction Results.

| Approach | 6-month Post Discharge |  |  | 1-year Post Discharge |  |  |
| --- | --- | --- | --- | --- | --- | --- |
| | wAUC | Trigram Count | Non-zero Trigram Count, $N(\%)$ | wAUC | Trigram Count | Non-zero Trigram Count, $N(\%)$ |
| Top-ngram | 0.905 | 200,000 | 199,712 (99.86%) | 0.852 | 200,000 | 199,771 (99.9%) |
| GatorTron Medium | 0.863 | 264,857 | 180,190 (68.03%) | 0.815 | 264,857 | 158,424 (59.85%) |

**Table 11:** Readmission Prediction Results.

| Approach | wAUC Score | Tetragram Count, $N$ | Non-zero Tetragram Count, $N(\%)$ |
| --- | --- | --- | --- |
| Top-ngram | 0.726 | 200,000 | 113,792 (56.9%) |
| BlueBERT | 0.725 | 208,455 | 40,611 (19.48%) |

### 6.2. Hospital Readmission Prediction

We extracted patients with obstructive sleep apnea (OSA) or atrial fibrillation (AF). Patients’ discharge notes were labeled as 1 if they were readmitted and labeled as 0 if not readmitted or deceased. The study cohort comprised a total of 67,161 patients, with 42,124 readmitted and 25,037 not readmitted.

Two approaches were explored. The Top-ngram approach used the top 200,000 most frequently appearing tetragrams in discharge notes, while the BlueBERT approach merged the top tetragrams from each health condition’s BlueBERT-expanded tetragram lists, resulting in a final tetragram count of 208,455.

Both the Top-ngram and BlueBERT approaches demonstrated similar predictive performance in readmission analysis, as indicated by nearly identical wAUC scores. Despite BlueBERT having a greater total number of tetragrams, only a smaller fraction contributed to the model. This suggests that the selection of tetragrams via BlueBERT produced a focused feature set, possibly resulting in more relevant but sparser features. In contrast, the n-gram method seems to benefit from a broader, denser feature utilization.

## 7. Discussions

Research suggests that obstructive sleep apnea increases the risk of cardiovascular and metabolic issues. Its frequent undiagnosed and delayed treatment lead to poor patient outcomes and substantial economic burdens. Deep phenotyping is crucial for early treatment, targeted interventions, improved patient outcomes, and more efficient healthcare resource allocation.

This study aims to utilize large language models (LLMs) and natural language processing to develop a lexicon specific to various OSA subphenotypes. This lexicon will help distinguish between patients diagnosed with OSA and its associated comorbidities, enhancing patient diagnosis and understanding of the condition and its related disorders.

### 7.1. Major Findings

The BioClinical BERT and BlueBERT outperformed all other tested models in diagnosing patients with OSA and heart failure. Both LLMs were pre-trained on one clinical dataset: MIMIC-III, potentially enabling the models to capture dataset-specific patterns and gain domain-specific knowledge, thereby improving their performance. Additionally, a more complex version of an LLM outperformed its simpler counterparts, even with fewer trigram features (Table 5 and Figure 2). This suggests that complex models are more adept at selecting higher-quality ngrams, leading to a more accurate characterization of health conditions.

Large language models can identify informative lexicons for OSA sub-phenotyping in simple patient cohorts, achieving wAUC scores of 0.9 or slightly higher (Table 8, Figure 2, and Figure 3). However, our preliminary study with more complex patient groups showed more challenges in effectively distinguishing between subphenotypes.

The Top-ngram approach for OSA sub-phenotyping, while yielding comparable or better results (Table 8), is more complex and computationally intensive compared to the models built with LLM-expanded lexicon. Although the LLM approach began with a larger pool of ngrams, only a smaller fraction had non-zero coefficients, indicating a simpler and interpretable model with a reduced risk of overfitting. This sparse modeling potentially contributes to the LLM approach’s speed, which is approximately five times faster than the Top-ngram approach. In contrast, the Top-ngram method utilizes a broader range of features, potentially leading to a denser feature set, as opposed to the focused and relevant feature set developed by LLMs.

The lexicons developed by large language models exhibited some utility, albeit limited, in predicting mortality risk and readmission. Both the Top-ngram and LLM approaches demonstrated comparable predictive performance, reflected in nearly identical wAUC scores. However, models generated from the LLM approach are simpler, a low percentage of non-zero features.

### 7.2. Limitations and Future Work

In this study, we explored the effectiveness of employing LLM-expanded lexicons for various sub-phenotyping of OSA and predicting patient outcomes. One limitation is that discharge notes for patients with OSA and comorbidities were labeled using ICD codes, primarily designed for billing and not necessarily indicative of the patient’s final diagnosis. To enhance model validation, we will consider incorporating two or more instances of diagnostic codes for a specific health condition when labeling (Keenan et al., 2020) or collaborating with physicians in this study to create a ground truth dataset (Cade et al., 2022).

Additionally, the ngram size study (Section 4.2) demonstrated that the bigram model generally out-performs the trigram and tetragram models. We will further investigate the impact of ngram size by applying it to phenotyping other patient cohorts, such as OSA and T2DM, OSA and HTN, and OSA and AF, as well as predicting patient mortality and hospital readmission risks.

Furthermore, the diagnostic and outcome prediction solely relied on unstructured clinical notes. Our future investigations will focus on integrating both structured data (such as demographics and time series measurements) and unstructured data (such as clinical notes) from EHRs for comprehensive phenotyping of OSA patients, as well as predicting mortality and readmission risks.

Other areas for future work include developing lexicons using a combination of ngram sizes, constructing models for complex patient cohorts, exploring advanced machine learning models beyond MLR, and delving into improved methods for explaining model outputs to benefit clinicians from our study.

## Data Availability

All data produced in the present study are available upon reasonable request to the authors.

https://physionet.org/content/mimiciv/2.2/

## Acknowledgments

We thank The National Energy Research Scientific Computing Center, The U.S. Department of Energy, The Sustainable Research Pathways Program, and The Hood College Volpe Scholarship for supporting the project. We are grateful to the physicians from Veterans Affairs for providing medical guidance.

## Notes

### Competing Interest Statement

The authors have declared no competing interest.

### Author Declarations

This paper used the MIMIC-IV dataset, which is available on the PhysioNet repository (https://physionet.org/content/mimiciv/2.2/).

## References

Emily Alsentzer, John R Murphy, Willie Boag, Wei-Hung Weng, Di Jin, Tristan Naumann, and Matthew McDermott. Publicly available clinical bert embeddings. arXiv preprint 1904.03323, 2019.

Awais Ashfaq, Anita Sant’Anna, Markus Lingman, and S-lawomir Nowaczyk. Readmission prediction using deep learning on electronic health records. Journal of biomedical informatics, 97:103256, 2019.

Adam V Benjafield, Najib T Ayas, Peter R Eastwood, Raphael Heinzer, Mary SM Ip, Mary J Morrell, Carlos M Nunez, Sanjay R Patel, Thomas Penzel, Jean-Louis Pépin, et al. Estimation of the global prevalence and burden of obstructive sleep apnoea: a literature-based analysis. Lancet Respir. Med., 7(8):687–698, August 2019.

Willie Boag, Dustin Doss, Tristan Naumann, and Peter Szolovits. What’s in a note? unpacking predictive value in clinical note representations. AMIA Summits on Translational Science Proceedings, 2018:26, 2018.

Rishi Bommasani, Drew A Hudson, Ehsan Adeli, Russ Altman, Simran Arora, Sydney von Arx, Michael S Bernstein, Jeannette Bohg, Antoine Bosselut, Emma Brunskill, et al. On the opportunities and risks of foundation models. 2108.07258, August 2021.

Brian E Cade, Syed Moin Hassan, Hassan S Dashti, Melissa Kiernan, Milena K Pavlova, Susan Redline, and Elizabeth W Karlson. Sleep apnea phenotyping and relationship to disease in a large clinical biobank. JAMIA Open, 5(1):ooab117, April 2022.

Youngjin Chae and Thomas Davidson. Large language models for text classification: From zeroshot learning to fine-tuning. Open Science Foundation, 2023.

Pei-Fu Chen, Lichin Chen, Yow-Kuan Lin, Guo-Hung Li, Feipei Lai, Cheng-Wei Lu, Chi-Yu Yang, Kuan-Chih Chen, and Lin Tzu-Yu. Predicting postoperative mortality with deep neural networks and natural language processing: model development and validation, 2022.

Munish Goyal and Jeremy Johnson. Obstructive sleep apnea diagnosis and management. Missouri medicine, 114(2):120, 2017.

Kexin Huang, Jaan Altosaar, and Rajesh Ranganath. Clinicalbert: Modeling clinical notes and predicting hospital readmission. arXiv preprint 1904.05342, 2019.

Mengqi Jin, Mohammad Taha Bahadori, Aaron Colak, Parminder Bhatia, Busra Celikkaya, Ram Bhakta, Selvan Senthivel, Mohammed Khalilia, Daniel Navarro, Borui Zhang, et al. Improving hospital mortality prediction with medical named entities and multimodal learning. arXiv preprint 1811.12276, 2018.

A. Johnson, L. Bulgarelli, and et al. L. Shen. MIMIC-IV, a freely accessible critical care database. Scientific Data, 10(1), 2023a. doi: 10.1038/s41597-022-01899-x.

A. Johnson, L. Bulgarelli, T. Pollard, S. Horng, L. A. Celi, and R. Mark. MIMIC-IV(version 2.2), 2023b.

Brendan T Keenan, H Lester Kirchner, Olivia J Veatch, Kenneth M Borthwick, Vicki A Davenport, John C Feemster, Maged Gendy, Thomas R Gossard, Frances M Pack, Laura Sirikulvadhana, et al. Multisite validation of a simple electronic health record algorithm for identifying diagnosed obstructive sleep apnea. J. Clin. Sleep Med., 16 (2):175–183, February 2020.

Swaraj Khadanga, Karan Aggarwal, Shafiq Joty, and Jaideep Srivastava. Using clinical notes with time series data for icu management. arXiv preprint 1909.09702, 2019.

Teven Le Scao, Angela Fan, Christopher Akiki, Ellie Pavlick, Suzana Ilić, Daniel Hesslow, Roman Castagné, Alexandra Sasha Luccioni, François Yvon, Matthias Gallé, et al. Bloom: A 176b-parameter open-access multilingual language model. eprint 2211.05100, 2022.

Bastien Lechat, Ganesh Naik, Amy Reynolds, Atqiya Aishah, Hannah Scott, Kelly A Loffler, Andrew Vakulin, Pierre Escourrou, R Doug McEvoy, Robert J Adams, et al. Multinight prevalence, variability, and diagnostic misclassification of obstructive sleep apnea. Am. J. Respir. Crit. Care Med., 205(5):563–569, March 2022.

Jinhyuk Lee, Wonjin Yoon, Sungdong Kim, Donghyeon Kim, Sunkyu Kim, Chan Ho So, and Jaewoo Kang. Biobert: a pre-trained biomedical language representation model for biomedical text mining. Bioinformatics, 36(4):1234–1240, 2020.

Yoon K Loke, J William L Brown, Chun Shing Kwok, Alagaratnam Niruban, and Phyo K Myint. Association of obstructive sleep apnea with risk of serious cardiovascular events: a systematic review and meta-analysis. Circ. Cardiovasc. Qual. Outcomes, 5(5):720–728, September 2012.

Renqian Luo, Liai Sun, Yingce Xia, Tao Qin, Sheng Zhang, Hoifung Poon, and Tie-Yan Liu. Biogpt: generative pre-trained transformer for biomedical text generation and mining. Briefings in Bioinformatics, 23(6):bbac409, 2022.

Michelle Olaithe, Romola S Bucks, David R Hillman, and Peter R Eastwood. Cognitive deficits in obstructive sleep apnea: Insights from a metareview and comparison with deficits observed in COPD,insomnia, and sleep deprivation. Sleep Med. Rev., 38:39–49, April 2018.

Joshua Parreco, Antonio Hidalgo, Robert Kozol, Nicholas Namias, and Rishi Rattan. Predicting mortality in the surgical intensive care unit using artificial intelligence and natural language processing of physician documentation. The American Surgeon, 84(7):1190–1194, 2018.

Sajan B Patel and Kyle Lam. Chatgpt: the future of discharge summaries? The Lancet Digital Health, 5(3):e107–e108, 2023.

Yifan Peng, Shankai Yan, and Zhiyong Lu. Transfer learning in biomedical natural language processing: an evaluation of bert and elmo on ten benchmarking datasets. arXiv preprint 1906.05474, 2019.

Javed Qadrud-Din, Ashraf Bah Rabiou, Ryan Walker, Ravi Soni, Martin Gajek, Gabriel Pack, and Akhil Rangaraj. Transformer based language models for similar text retrieval and ranking. arXiv preprint 2005.04588, 2020.

Jayroop Ramesh, Niha Keeran, Assim Sagahyroon, and Fadi Aloul. Towards validating the effectiveness of obstructive sleep apnea classification from electronic health records using machine learning. In Healthcare, volume 9, page 1450. MDPI, 2021.

Winfried J Randerath, Simon Herkenrath, Marcel Treml, Ludger Grote, Jan Hedner, Maria Rosaria Bonsignore, Jean Louis Pépin, Silke Ryan, Sophia Schiza, Johan Verbraecken, et al. Evaluation of a multicomponent grading system for obstructive sleep apnoea: the baveno classification. ERJ Open Research, 7(1), 2021.

Peter N Robinson. Deep phenotyping for precision medicine. Hum. Mutat., 33(5):777–780, May 2012.

Jaromir Savelka and Kevin D Ashley. The unreasonable effectiveness of large language models in zeroshot semantic annotation of legal texts. Frontiers in Artificial Intelligence, 6, 2023.

Andrea Seiler, Millene Camilo, Lyudmila Korostovtseva, Alan G Haynes, Anne-Kathrin Brill, Thomas Horvath, Matthias Egger, and Claudio L Bassetti. Prevalence of sleep-disordered breathing after stroke and TIA: A meta-analysis. Neurology, 92(7): e648–e654, February 2019.

Karan Singhal, Shekoofeh Azizi, Tao Tu, S Sara Mahdavi, Jason Wei, Hyung Won Chung, Nathan Scales, Ajay Tanwani, Heather Cole-Lewis, Stephen Pfohl, et al. Large language models encode clinical knowledge. Nature, 620(7972):172–180, 2023.

Satu Strausz, Sanni Ruotsalainen, Hanna M Ollila, Juha Karjalainen, Tuomo Kiiskinen, Mary Reeve, Mitja Kurki, Nina Mars, Aki S Havulinna, Elina Luonsi, et al. Genetic analysis of obstructive sleep apnoea discovers a strong association with cardiometabolic health. bioRxiv, 2020. doi: 10.1101/2020.08.04.235994.

Loreto Sulit, Amy Storfer-Isser, H Lester Kirchner, and Susan Redline. Differences in polysomnography predictors for hypertension and impaired glucose tolerance. Sleep, 29(6):777–783, June 2006.

Nathaniel F Watson. Health care savings: The economic value of diagnostic and therapeutic care for obstructive sleep apnea. J. Clin. Sleep Med., 12 (08):1075–1077, August 2016.

Jun Wu, Xuesong Ye, Chengjie Mou, and Weinan Dai. Fineehr: Refine clinical note representations to improve mortality prediction. In 2023 11th International Symposium on Digital Forensics and Security (ISDFS), pages 1–6. IEEE, 2023.

Wanyuan Xia, Yanhong Huang, Bin Peng, Xin Zhang, Qingmeng Wu, Yiying Sang, Yetao Luo, Xun Liu, Qian Chen, and Kaocong Tian. Relationship between obstructive sleep apnoea syndrome and essential hypertension: a dose-response meta-analysis. Sleep Med., 47:11–18, July 2018.

Kristine Yaffe, Alison M. Laffan, Stephanie Litwack Harrison, Susan Redline, Adam P. Spira, Kristine E. Ensrud, Sonia Ancoli-Israel, and Katie L. Stone. Sleep-Disordered Breathing, Hypoxia, and Risk of Mild Cognitive Impairment and Dementia in Older Women. JAMA, 306(6):613–619, 08 2011. ISSN 0098-7484. doi: 10.1001/jama.2011.1115.

Xi Yang, Aokun Chen, Nima PourNejatian, Hoo Chang Shin, Kaleb E Smith, Christopher Parisien, Colin Compas, Cheryl Martin, Anthony B Costa, Mona G Flores, et al. A large language model for electronic health records. NPJ Digital Medicine, 5(1):194, 2022.

Jiancheng Ye, Liang Yao, Jiahong Shen, Rethavathi Janarthanam, and Yuan Luo. Predicting mortality in critically ill patients with diabetes using machine learning and clinical notes. BMC medical informatics and decision making, 20(11):1–7, 2020.

